# The Portland SEEQ: A Harmonizable Questionnaire for Environmental Exposome Profiling in Prodromal Synucleinopathy

**DOI:** 10.64898/2026.10.26.26366216

**Authors:** Lee E. Neilson, Jessica Locke, Hannah Cunningham, Jonathan E. Elliott, Miranda M. Lim

## Abstract

**Objective:** To describe development of the Portland Suspected Environmental Exposures Questionnaire (SEEQ) and its initial application for environmental exposome profiling in isolated REM sleep behavior disorder (iRBD).

**Background:** Environmental exposures may contribute to Parkinson disease and related synucleinopathies, yet existing instruments variably capture specific agents, co-exposures, exposure uncertainty, timing, and mitigating factors.

**Methods:** SEEQ was iteratively developed to assess pesticides, solvents/industrial chemicals, welding fumes, heavy metals, residential and occupational exposures, and relevant lifestyle factors. Direct exposures were classified as definite, probable, possible, or uncertain/unlikely, with additional assessment of frequency/duration, life stage, and protective-equipment use. SEEQ was administered to participants with polysomnography-confirmed iRBD. Exposure prevalence, co-exposure patterns, and exploratory associations with clinical phenotypes and longitudinal progression were examined.

**Results:** Eighty-three participants completed SEEQ; 81 (97.6%) completed all four direct exposure domains. Definite/probable exposure was reported for pesticides in 38.6%, solvents/industrial chemicals in 43.4%, welding fumes in 21.0%, and heavy metals in 34.1%. Among complete responders, 60.5% reported ≥1 and 42.0% ≥2 direct exposure domains. Forty-five participants identified ≥1 named agent or mixture. Greater exposure burden was cross-sectionally associated most consistently with greater autonomic symptom burden, but was not associated with longitudinal clinical worsening.

**Conclusion:** SEEQ provides a practical, multidomain framework for structured environmental exposome profiling in prodromal synucleinopathy. Larger studies with external exposure validation and longitudinal follow-up are needed.

## Introduction

Parkinson’s Disease (PD) is the fastest growing neurological disorder worldwide, outpacing the increases expected due to aging and improved diagnosis^1^. Increasing attention is being paid to environmental^2^ and military-specific factors^3,4^. Most studies have investigated the link between a single exposure and PD incidence. However, the exposure histories of individual patients are often uncertain, multidimensional and remote from clinical assessment. In fact, a Movement Disorder Society task force has recently called for the need for a polyexposure score in PD^5^. This score could determine the potential influence of the exposome – acute and chronic external exposures, lifestyle factors, social interactions, and the endogenous biological processes which respond to these accumulated exposures. And, it would complement the growing evidence base about the genome.

Many tools exist for examining environmental exposures in PD^6-10^. However, these tools either require expert trained interviewers, only capture exposures at the category level, do not assess for personal protective equipment as a mitigating factor, and/or lack the nuance of uncertainty inherent to recall-based surveys. Moreover, none of these instruments have a dedicated military exposures module. To close this gap, we developed the Portland Suspected Environmental Exposures Questionnaire (SEEQ). Herein, we describe its design and initial implementation. We then deployed this survey in a cohort of people with idiopathic REM sleep behavior disorder (iRBD), to identify exposure yield and co-exposure patterns, and explore associations with clinical phenotypes and symptom progression.

## Methods

### SEEQ development

We performed an extensive literature review to understand the landscape of current questionnaires and emerging environmental risk factors for synucleinopathies. We then designed the first version of the survey to capture the relevant exposure domains with additional information on dose, timing, and mitigating factors, and to allow for some measure of uncertainty in recall. SEEQ was intentionally structured to permit hierarchical harmonization: detailed agent-level, certainty, timing, and PPE data can be retained for SEEQ-specific analyses, while responses can be collapsed to broader pesticide, solvent, welding, metal, residential, occupational, and lifestyle constructs compatible with established PD environmental questionnaires. Later revisions added plain-language examples to aid recognition, refined wording and skip logic, distinguished unidentified exposures from named but unlisted agents, and standardized selected lifestyle response conventions. No test–retest reliability, psychometric validation, criterion validation or biomarker confirmation was established.

Four direct domains address pesticides, solvents/industrial chemicals, welding fumes and heavy metals. Responses distinguish definite, probable, possible and uncertain/unlikely exposure. Follow-up items address frequency/duration, developmental timing and protective equipment where applicable, with recognition-based lists of pesticides, chemicals and metals. Additional modules record type of residence, well-water use, commercial-farm and Superfund proximity, occupational history, NSAIDs, caffeine, tobacco/vaping, physical activity and head injury. These contextual and lifestyle variables are not all treated as toxicant exposures, but are known to be relevant to synuclein biology.

### Exposure definitions and descriptive outcomes

Each qualifying exposure domain contributed one point. Broad exposure burden summed definite/probable pesticide, solvent/industrial chemical, welding-fume and heavy-metal exposure plus affirmative well-water use, commercial-farm proximity and Superfund proximity responses (range 0–7). Regular exposure burden counted the four direct domains only when exposure was definite/probable and frequency/duration was “Regularly for <1 year” or “Regularly for 1+ years” (range 0–4). Possible or uncertain responses and blank frequency responses earned zero. Any missing required domain responses prevented scoring.

### Study participants

Individuals with iRBD at our local institution were invited to participate as part of a larger, ongoing observational cohort study. To be eligible, iRBD diagnosis requires confirmation of REM sleep without atonia (RSWA) by in-lab video PSG according to American Academy of Sleep Medicine guidelines with at least the minimal six channel electroencephalography montage, electrooculography of both eyes, and electromyography (EMG) of at least chin and two legs. The presence of RSWA in addition to either clinical history or video capture of dream enactment behavior constitutes an official diagnosis. Individuals with RBD and co-morbid narcolepsy are excluded from enrollment due to a known pathophysiology causing RBD that is unrelated to synucleinopathy. Each participant undergoes a comprehensive annual battery of assessments. For greater detail, see our protocol and baseline characteristics papers.

### Exploratory statistical analysis

Continuous variables are reported as median (IQR), with explicit denominators and missing counts. Baseline associations used unadjusted Spearman correlations and two-sided permutation p-values from 20,000 permutations with a +1 correction. Participant percentile-bootstrap intervals used 10,000 resamples for the focused baseline figure. Benjamini–Hochberg FDR was applied across the 12 focused comparisons (six measures by two indices). These outcomes were selected after a wider exploratory screen of 742 correlations; focused q-values are therefore not confirmatory and are reported alongside the wider-screen interpretation.

For participants with repeated measurements, an annual slope was estimated by ordinary least squares using all valid values and elapsed years calculated from visit dates (days/365.25). At least two valid measurements on distinct dates were required. MoCA and BSIT slopes were sign-reversed so positive slopes denoted worsening, matching increasing motor, autonomic and Trail B scores. Spearman correlations related exposure counts to these slopes; 3,000 participant bootstrap resamples provided confidence intervals and FDR covered ten primary tests. This equally weighted slope analysis does not model unequal slope precision or nonlinear change.

Agent-specific and pesticide-definition comparisons used separate OLS models adjusted for baseline age, sex and education. HC3 robust standard errors and restricted/unrestricted wild-bootstrap-t procedures (19,999 Rademacher draws) provided p-values/95% intervals. FDR was applied within each exploratory family: seven Agent Orange, seven fire/burn-pit, seven ten-agent and 28 pesticide-definition comparisons. The pesticide definitions were any suspected; definite/probable plus regular duration; definite plus regular duration ≥1 year; and that sustained category plus reported exposure without PPE. The last definition uses a yes/no PPE item, not a direct measure of how regularly PPE was omitted. Missing outcomes were not imputed.

### Ethics

All participants signed informed consent. The institutional review board of OHSU gave ethical approval for this work.

## Results

### Participants and response availability

A total of 83 consecutive participants with iRBD completed the SEEQ, with 51 having a minimum of 2 comprehensive visits, separated 1 year apart (**Table 1**). Baseline median age was 69.0 years (IQR 57.5–72.0), 16.9% were female, and median education was 16.0 years (14.0–17.0). All four direct domains were completed by 81/83 (97.6%) participants. Forty-five participants (54.2%) identified at least one named agent or mixture. Eighteen had documented unidentified pesticide exposure.

**Table 1.** Cohort characteristics and questionnaire response availability.

| **Characteristic** | **Summary Statistic** |
| --- | --- |
| **Demographics** |  |
| Age, years | 69.0 (57.5–72.0) |
| Sex, n (%) female | 10 (16.9%) |
| Education, years | 16.0 (14.0–17.0) |
| **RBD / clinical characteristics** |  |
| Age at RBD symptom onset, years | 52.5 (33.0–63.2) |
| RBD symptom duration, years<sup>1</sup> | 10.0 (4.8–23.2) |
| MoCA total | 26.0 (23.0–28.0) |
| MDS-UPDRS Part III total | 4.0 (2.0–7.8) |
| SCOPA-AUT total | 16.0 (8.5–23.0) |
| BSIT total | 8.0 (5.5–10.0) |
| Trail Making Test B, seconds | 79.0 (60.0–103.0) |
| **SEEQ feasibility / completion** |  |
| Participants completing SEEQ, n<sup>3</sup> | 83 |
| Participants identifying ≥1 named agent, n (%)□ | 45 (54.2%) |
| Participants reporting unknown/unidentified pesticide exposure, n (%)□ | 18 (45.0%) |
| **Direct exposure prevalence — definite/probable** |  |
| Any direct exposure domain, n (%) | 49 (60.5%) |
| Pesticides, n (%) | 32 (38.6%) |
| Solvents/industrial chemicals, n (%) | 36 (43.4%) |
| Welding fumes, n (%) | 17 (21.0%) |
| Heavy metals, n (%) | 28 (34.1%) |
| **Number of definite/probable direct exposure domains** |  |
| 0 domains, n (%) | 32 (39.5%) |
| 1 domain, n (%) | 15 (18.5%) |
| 2 domains, n (%) | 11 (13.6%) |
| 3 domains, n (%) | 17 (21.0%) |
| 4 domains, n (%) | 6 (7.4%) |
| Median number of direct exposure domains (IQR) | 1 (0–3) |
| **Other exposure-context variables** |  |
| Well-water use, n (%) | 26 (31.3%) |
| Lived near commercial farm, n (%) | 16 (19.3%) |
| Lived near Superfund site, n (%) | 5 (6.0%) |
| Any prespecified high-risk occupation, n (%)□ | 66 (79.5%) |

Continuous values are median (IQR); percentages use available responses unless the row describes status in the full cohort. Missing percentages use N=83 and include absent clinical linkage. Completion denotes retained questionnaire records. Duration is baseline age minus onset age. Unidentified-agent fields are incompletely populated; the single solvent affirmative is not an interpretable cohort prevalence. Occupation reflects any of the 18 listed contexts, not validated risk. Phenoconversion is provisional first recorded follow-up diagnosis. Uncertain contextual responses are available but not affirmative.

### Exposure prevalence and co exposure

Definite/probable exposure was reported by 32/83 (38.6%) for pesticides, 36/83 (43.4%) for solvents/industrial chemicals, 17/81 (21.0%) for welding fumes and 28/82 (34.1%) for heavy metals. Among 81 complete responders, 49 (60.5%) reported at least one domain and 34 (42.0%) at least two. The distribution was 32 with zero, 15 with one, 11 with two, 17 with three and six with four domains; median count was 1 (0–3). Regular direct-domain counts were zero in 40, one in 21, two in 12, three in seven and four in one participant (**Figure 1**). A simultaneous depiction of both the prevalence of individual exposure domains and the patterns of multidomain co-exposure is shown in **Figure 3**.

**Figure 1.**
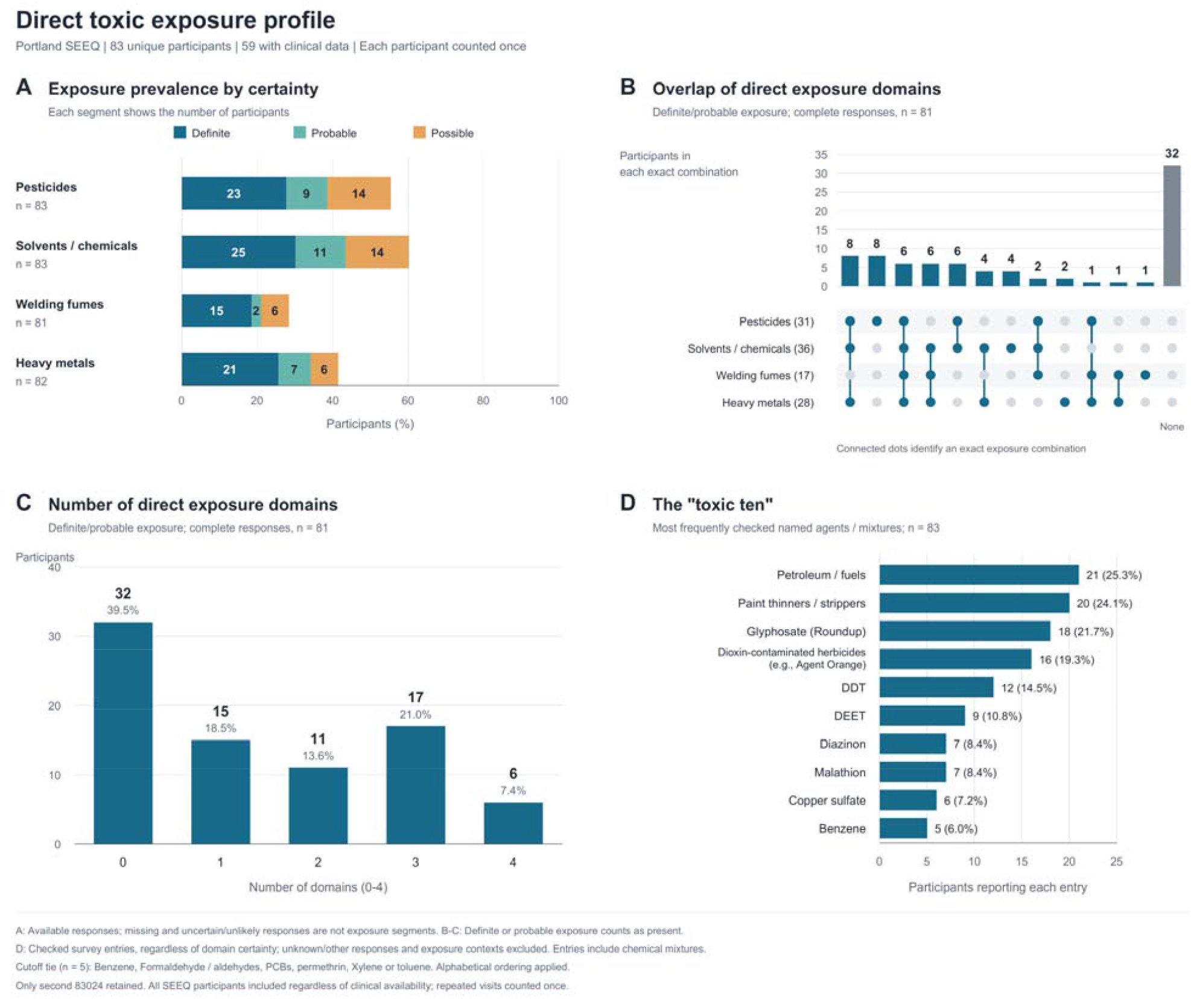
Direct exposure profile. A: definite/probable/possible prevalence with domain-specific denominators. B: exact intersections of definite/probable domains among 81 complete responders. C: number of domains among those 81. D: ten most frequently checked named chemicals/mixtures among 83. Generic contexts and unknown/other entries are excluded. Ties at five reports were resolved alphabetically; PCBs and other tied entries may lie outside the displayed ten. This prevalence ranking differs from the selected ten-agent hypothesis panel in Figure 3.

A total of 53 of 83 respondents (63.9%) reported military service. Of these, 15 (28.3%) reported dioxin-contaminated herbicide exposure during military service, including Agent Orange. Distribution of military-related exposures is reported in **Table 2**.

**Table 2.** Survey responses to military service module.

| **Reported exposure** | **n (%)** |
| --- | --- |
| Dioxin-contaminated herbicides, including Agent Orange | 15 (28.3%) |
| Chemical fires or burn pits | 12 (22.6%) |
| Both Agent Orange and chemical fires/burn pits | 5 (9.4%) |
| Either Agent Orange or chemical fires/burn pits | 22 (41.5%) |
| Trichloroethylene (TCE) | 2 (3.8%) |
| Tetrachloroethylene (PCE/PERC) | 4 (7.5%) |

### Exploratory clinical associations

Broad and regular exposure counts correlated with higher baseline SCOPA-AUT scores (ρ=0.431, p<0.001; ρ=0.348, p<0.01; **Figure 2**). Broad exposure also correlated nominally with lower MoCA (ρ=−0.289, p=0.03), while the other associations were directionally concordant. However, regular burden may be underestimated. In the clinical cohort, qualifying exposure lacked frequency in ten welding, one heavy-metal and one solvent response, affecting 11 participants overall.

**Figure 2.**
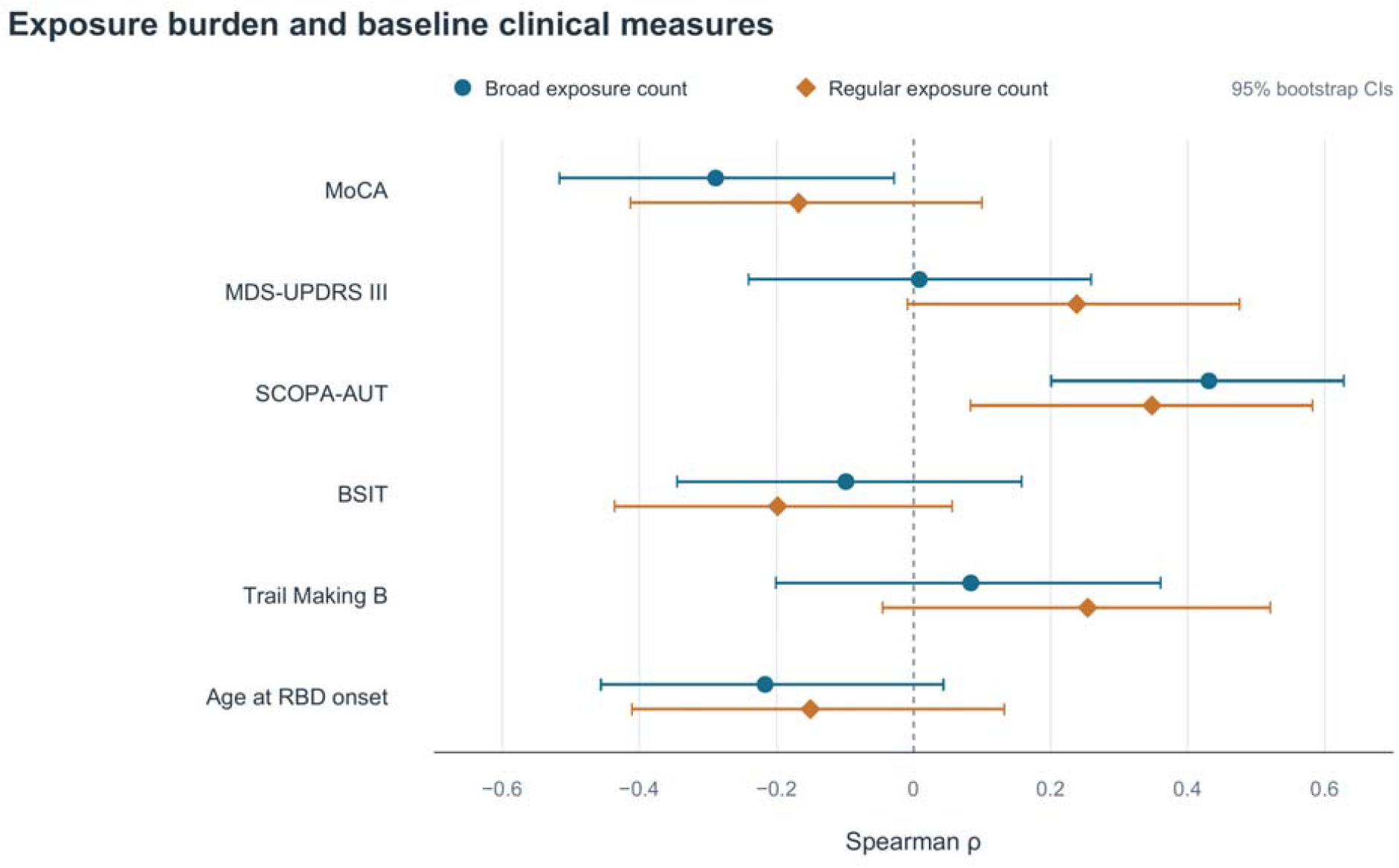
Broad and regular exposure counts and baseline clinical measures. Unadjusted Spearman correlations, original score directions. Intervals are marginal 95% percentile-bootstrap CIs from 10,000 participant resamples. Pairwise n=59,58,59,59,45,56 in displayed order. Focused FDR covers 12 tests and does not replace the broader 742-test correction. Greater MoCA/BSIT values indicate better performance; greater motor/autonomic/Trail B values indicate worse symptoms/performance.

**Figure 3.**
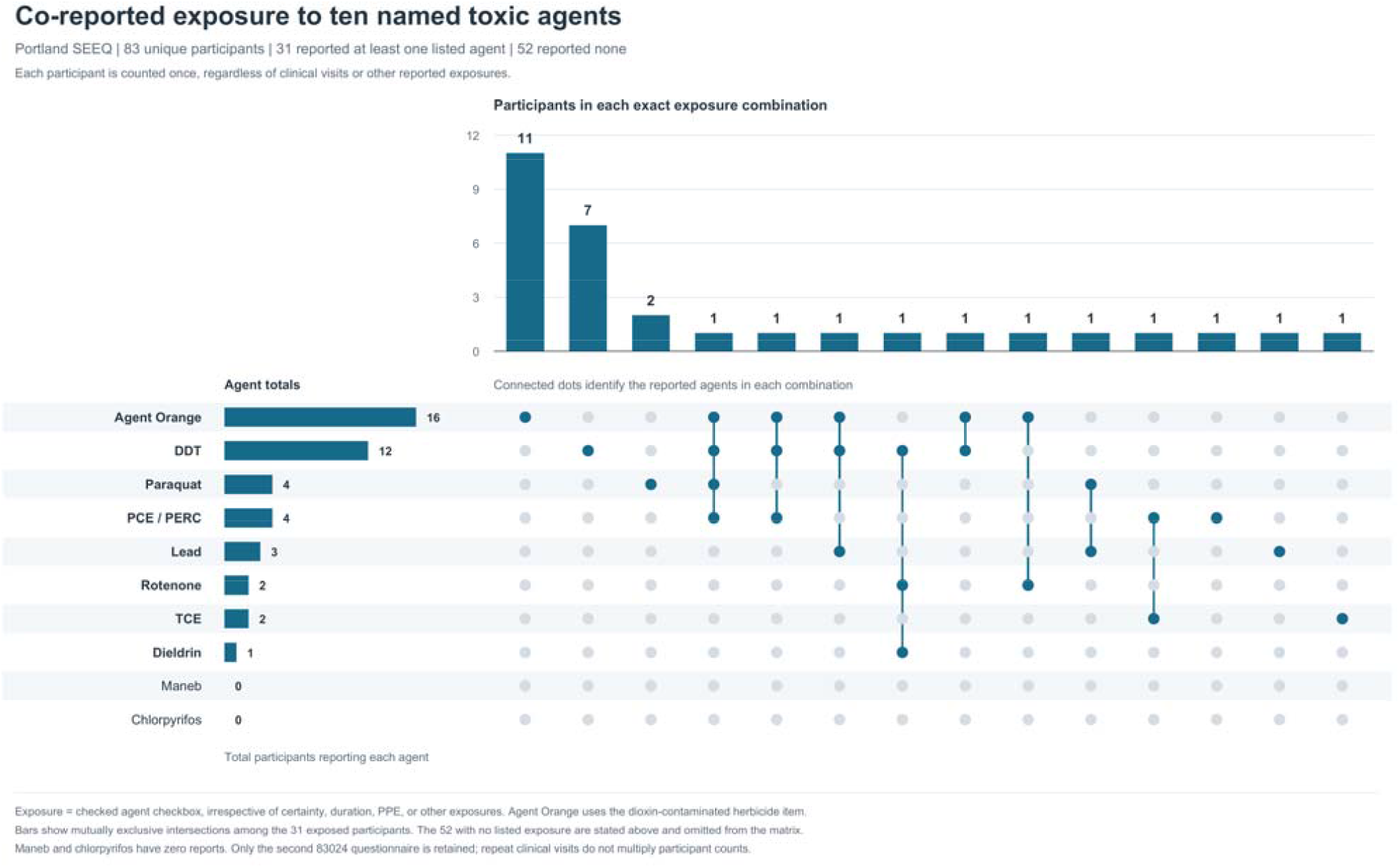
Co reported exposure to ten selected agents. Horizontal bars show marginal reporting counts; vertical bars show mutually exclusive agent intersections. Thirty-one of 83 reported at least one selected agent, with 14 nonempty combinations. Fifty-two reported none and are omitted from the matrix. Maneb and chlorpyrifos are retained as zero-count rows. Agent Orange uses the dioxin-contaminated herbicide item. Other exposures, frequency and certainty do not change membership.

Among 51 participants with repeated visits, median follow-up was 2.89 years (1.90–3.86). No exposure-count association with annual worsening was nominally significant. Thirteen participants reporting Agent Orange and 46 not reporting it had baseline clinical data. Unadjusted comparisons suggested differences in motor function and executive function measures (**Figure 4**).

**Figure 4.**
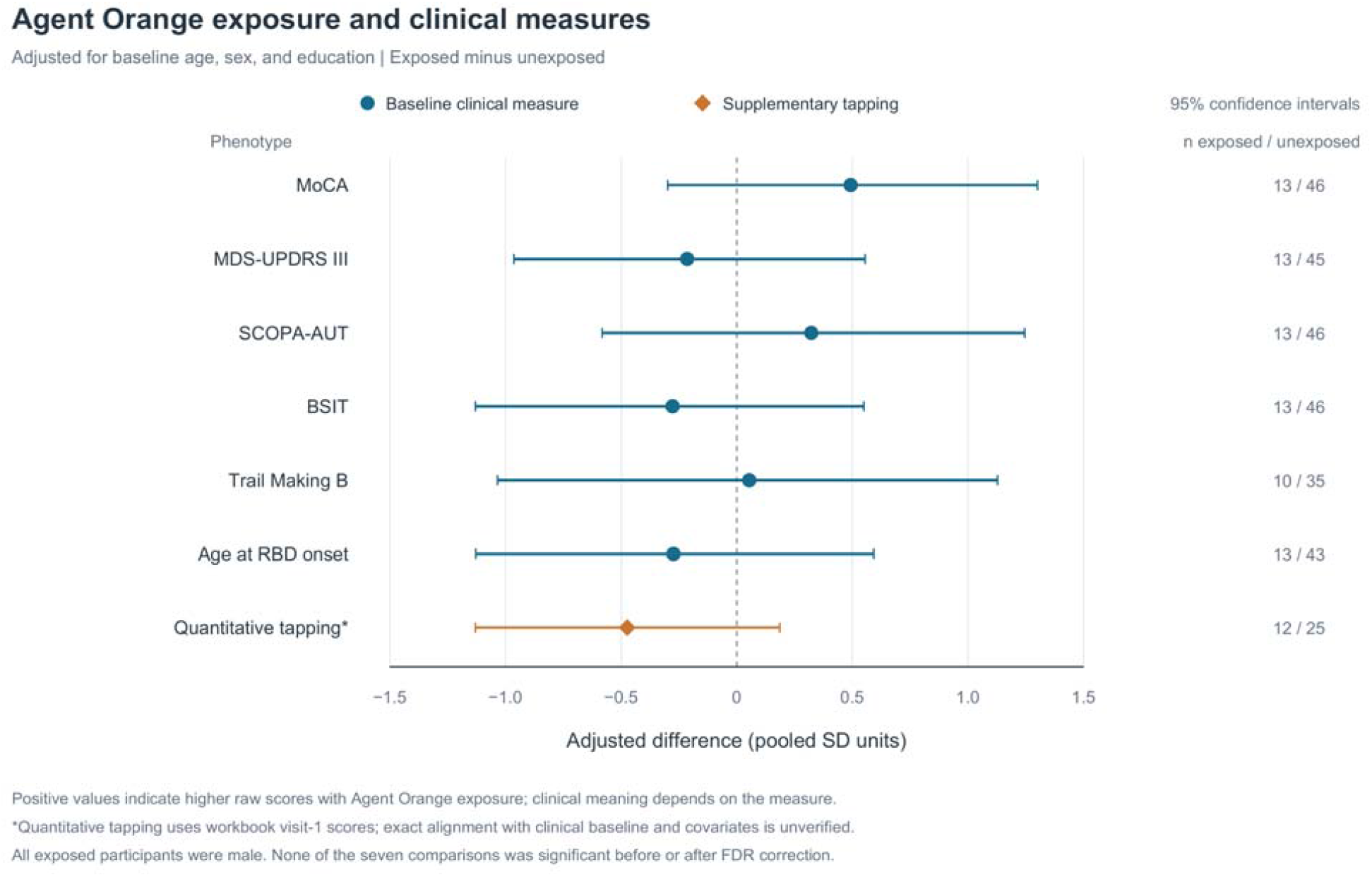
Agent Orange standardized adjusted differences.

## Discussion

Herein, we report the initial development and deployment of the Portland SEEQ within a population of individuals with iRBD. Survey completion rates were high and the participants reflected the larger pattern of demographics seen in studies of iRBD^11,12^. Direct-domain response availability was high, and exposure patterns were heterogeneous. More than two-fifths of complete responders reported at least two definite/probable direct domains. The named-agent panel additionally demonstrated that single-agent and overlapping reports coexist, supporting analysis of exposure profiles rather than assuming one dominant exposure per participant. We also show that those with greater exposure burdens, either collectively or with single agents, show a consistent directional trend of association with greater symptom burden. The pilot nature of this demands we interpret these associations cautiously.

A useful feature of the questionnaire is its preservation of uncertainty. Definite, probable and possible reports can be analyzed separately or collapsed under explicit rules; frequency and PPE provide additional dimensions without implying that participants can reconstruct historical dose accurately. Recognition prompts and agent examples may support recall, but improved recall and reduced cognitive burden have not been demonstrated. This is likely to reduce intra-rater variability, though this was not formally tested. Equal-weight broad and regular counts offer reproducible summaries, yet combine exposures with different timing, intensity and biological plausibility. They should not be interpreted as equivalent toxic doses and future work to develop a polyexposure score is needed.

As with survey-based measured, there is the possibility of recall bias, especially since this is a diseased population, and not compared against a control population. Likewise, there is no direct biochemical measurement which can provide objective evidence for exposure. Of the existing instruments, only one is considered formally validated despite the absence of a formal validation paper^6^ and a second does have some external exposure validation but not formal questionnaire validation: pesticide information is mapped to pesticide product databases and both residential and workplace exposure can be reconstructed using California pesticide-use records geographic information systems. Future work with more specific validation measures – imaging, geocoding, using job exposure matrices, and tissue or biofluid analyses – is warranted. At this time, SEEQ is not intended to replace detailed industrial-hygiene exposure reconstruction or established standardized epidemiologic instruments. Rather, it is intended to provide a feasible first-line exposome phenotype that can identify plausible lifetime exposure patterns, quantify multidomain co-exposure, support harmonized pooled analyses, and nominate participants or exposures for deeper validation.

In conclusion, the Portland SEEQ provided structured self-reported environmental exposure profiles in a sample of people with iRBD. Clinical associations remain exploratory; prospective measurement and external validation are required before clinical or causal applications.

## Declarations

### Author contributions

MML: writing - review & editing

HC: formal analysis, writing - review & editing

JL: formal analysis, writing - review & editing

JEE: writing - review & editing

LEN: conceptualization, investigation, formal analysis, writing - original draft

All authors directly accessed and verified the underlying data reported in the manuscript, contributed to the revision of the manuscript, and read and approved its final version.

## Acknowledgments

None

## Data and materials availability

The data that support the findings of this study are available on request from the corresponding author. The data are not publicly available due to privacy or ethical restrictions.

